# SARS-CoV-2 detection in wastewater as an early warning: the case of metropolitan area of the city of Buenos Aires (AMBA)

**DOI:** 10.1101/2022.04.23.22273730

**Authors:** Alejandro Barrio, Verónica Borro, Marcelo Cicchino, Adriana Morón, Lorena Coronel, Juan Vuolo, Paula Mayón, Ayelén Moroz, Paula Maisa, Sebastián Alcántara, María Torras, Marcos Cervini, Martín Arzeno, Cristian Godoy, Diego Campillay, Nestor Filiel, Ana Salas, Paula Salvio

## Abstract

Agua y Saneamientos Argentinos S.A. (AySA) delivers essential services as drinking water production and wastewater treatment for more than 14.5 million inhabitants of Buenos Aires Metropolitan Area (AMBA), Argentina, and collects residual liquids of 8.5 million through 16,178 km network. Since the very moment the World Health Organization (WHO) declared the COVID-19 pandemic, AySA developed a methodology to determine SARS-CoV-2 viral genetic load in untreated wastewater as an epidemiological surveillance tool, based on international experiences. In order to monitor viral load in the representative areas of the sewage collection system, more than 1500 samples where concentrated, by using an adapted ultracentrifugation method followed by RNA extraction and quantitative reverse transcription polymerase chain reaction (RT-qPCR), to measure the target Orf1ab gene of SARS-CoV-2 in our molecular microbiology laboratory.

This research was developed for a period that lasted from January 01 to June 02, 2021, in order to anticipate to the current second wave. The results achieved have demonstrated that changes in SARS-CoV-2 RNA are satisfactorily related to local epidemiological data for COVID-19. The association of variables is statistically significant when analyzing data from four large wastewater treatment plants, (R2 > 0.5 and p-value < 0.05), obtaining significant correlations between log10 viral genomic load and log10 positive cases reported one and two weeks later after samples were analyzed.

From the results obtained, it is concluded that the virus sewage system levels were a good predictor of clinical cases to be diagnosed in the immediate future and it is feasible to use this methodology, at local level, as an additional tool for decision-making in public health strategy.

**Highlights:**

⍰ Development of an integrated molecular sampling, analysis and detection system that can alert about the circulation of SARS-CoV-2.
⍰ Early warning of infected population.
⍰ First laboratory in a public water company to develop a monitoring method in Argentina.
⍰ Use of poly aluminum chloride (PAC) as a coagulant in the viral concentration stage.

## 1. Introduction

At the beginning of 2020 the COVID-19 epidemic was declared by WHO as a public health emergency of international concern and, on March 11th, it was characterized as a pandemic [1], developing different strategies for the epidemiological monitoring of the infection took greater relevance. efore the first cases of infected patients were registered in the Netherlands, scientists from the KWR Water Research Institute [2] had already detected traces of coronavirus in wastewater corroborating the fact that, although SARS -CoV-2 is mainly an airborne virus, it is also excreted in feces and urine from symptomatic and asymptomatic patients [3–5]. These preliminary studies were followed by others countries, (for example Spain [6]) and thereby controlling SARS-CoV-2 in wastewater was quickly transferred to other as a monitoring tool and an epidemiological surveillance system.

Agua y Saneamientos Argentinos S.A. (AySA) provides drinking water and sanitation services for more than 14.5 million inhabitants of Buenos Aires Metropolitan Area (AMBA) and collects wastewater of 8.5 million through 16,178 km network.

In order to determine the viral genetic load of SARS-CoV-2 in untreated wastewater as a possible epidemiological surveillance tool, Agua y Saneamientos Argentinos S.A. (AySA), initially with the technical assistance of the Entidad Regional de Saneamiento y Depuración de Aguas Residuales (ESAMUR) [7], undertook the commitment as a response to the global trend of water and sanitation operators; working in parallel different academic and scientific institutions, whose common primary purpose was to assist government entities with information for decision-making in matters of public health.

The result of this initiative was the design of a sampling plan which included more than 1500 samples since May 2020 of representative areas of the sewage collection system and treatment plants, to assess the viral load by using an adapted ultracentrifugation technique as a concentration method, followed by RNA extraction and quantitative reverse transcription polymerase chain reaction (RT-qPCR) for measuring the Orf1ab gene target for SARS-CoV2 [8, 9] in our molecular microbiology laboratory.

## 2. Materials and Methods

### 2.1. Sampling sites and samples collection

The present study covers the period from December 2020 to June 2021. Weekly samples were taken from influents of four wastewater treatment plants Figure 1.

**Figure 1.**
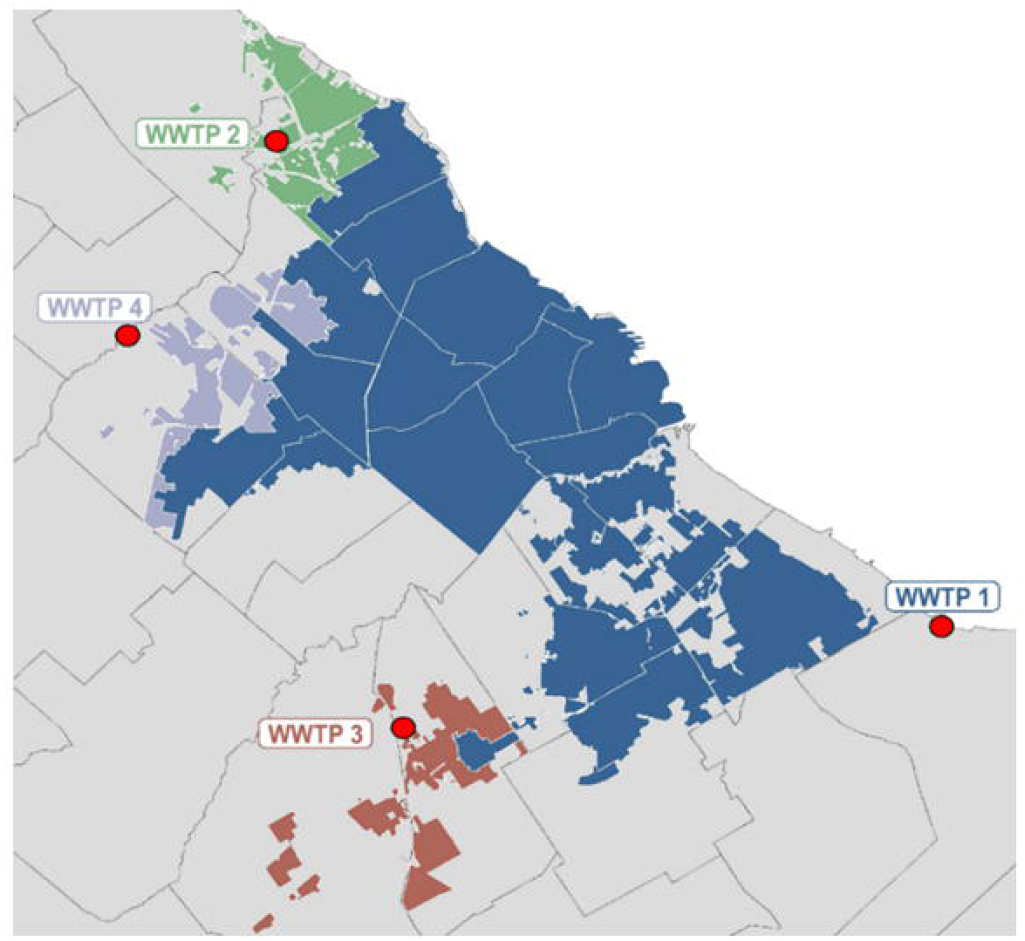
Map with the location of WWTP’s analysed in the study.

From each sample point, 3 liters of punctual samples were taken between 9 to 12 AM. These samples were divided in 1 liter for molecular analysis and 2 liters for additional determinationsTable 1. Samples were transported and preserved at 4 °C to AySA Central Laboratory until their concentration step.

**Table 1.** Characteristics of WWTP’s analysed: This table shows influent parameters for each treatment plant. Q indicates average flows calculated as the average of flows of the day’s sampled for the study

| WWTP (ID) | Q * (m<sup>3</sup>/day) | BOD5 (mg/l) | QOD (mg/l) | TSS (mg/l) | Total HC (mg/l) | Oil and Grease (mg/l) | SAAM (mg/l) | Phenolic Compounds (mg/l) | pH in situ |
| --- | --- | --- | --- | --- | --- | --- | --- | --- | --- |
| WWTP 1 | 1,895,900 | 112 | 237 | 99 | < 4 | 30 | 1.12 | 0.02 | 7.48 |
| WWTP 2 | 125,600 | 77 | 199 | 84 | < 4 | 18 | 0.63 | 0.02 | 6.67 |
| WWTP 3 | 36,200 | 142 | 469 | 272 | < 4 | 38 | 0.71 | 0.02 | 7.78 |
| WWTP 4 | 19,900 | 102 | 315 | 141 | < 4 | 26 | 1.20 | 0.03 | 7.86 |

Additionally, Escherichia coli concentrations were measured in tributaries of treatment plants, in order to identify the peak hours of tipping activity in the sewage system.Figure 2.

**Figure 2.**
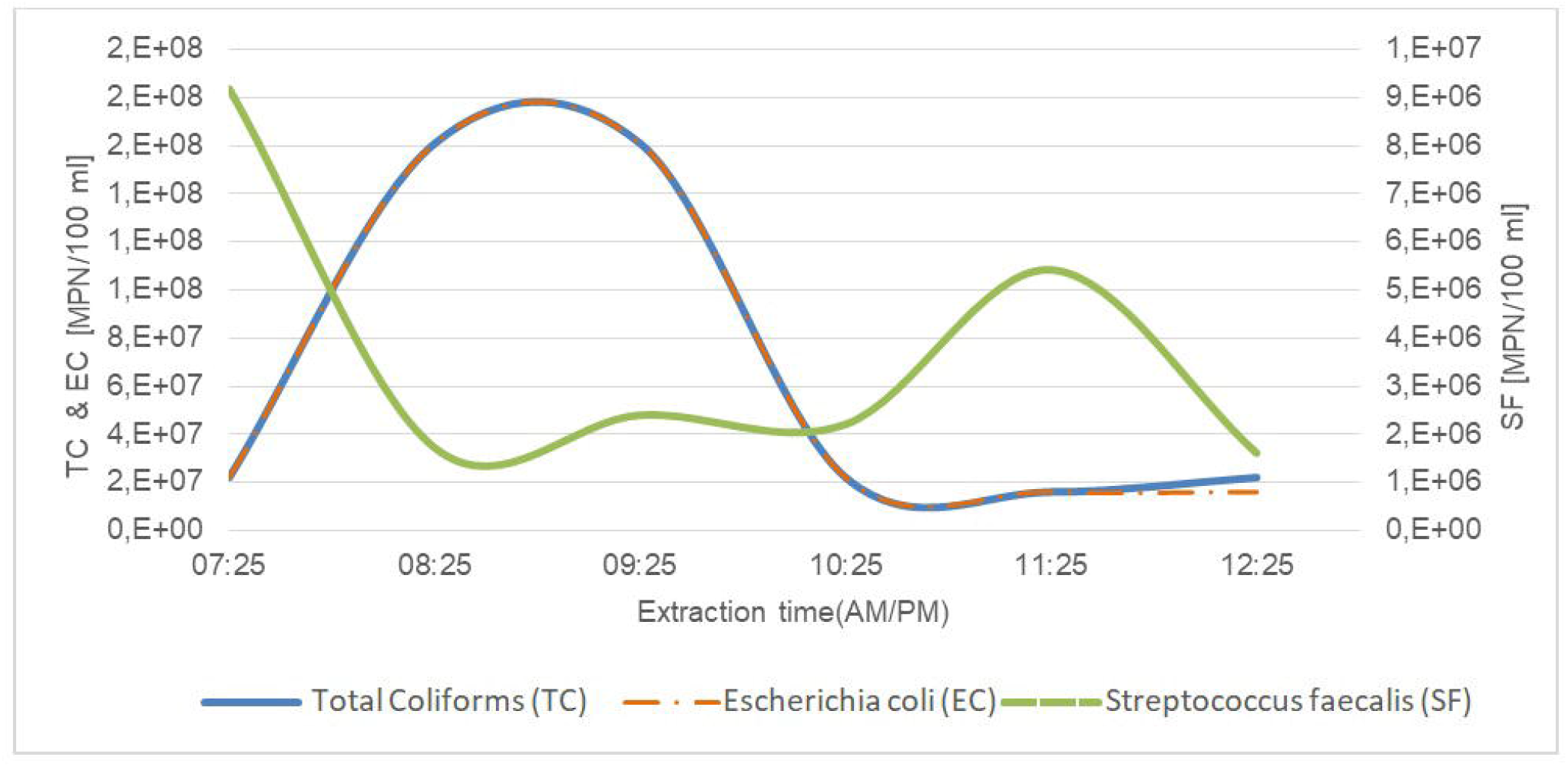
Colimetry Graph Example (WWTP 1).

### 2.2. Wastewater concentration

The samples were concentrated on the same day of the extraction.

The concentration process consisted of an ultracentrifugation adapted method. Briefly, an aliquot of 350 ml was taken from the homogenized sample and 3.5 ml of poly aluminum chloride (PAC) solution (0.9% m/V in aluminum) were added in a 500 ml centrifuge tube (Corning - ref 431123). The mixture was homogenized by vortex (Vortexer) and pH was adjusted to 6.0 with 1 N HCl or 2.5 N NaOH. Then a centrifugation at 1700 x g 20 min at 15 °C was made.

The supernatant was discarded and the pellet was resuspended with 8.75 ml of 3% m/v meat extract solution (Merck) by shaking at 1000 rpm, 1 minute. A second centrifugation at 1900 x g, 30 min at 15°C was made and supernatant was discarded again. The new pellet was resuspended in 1.75 ml of PBS and homogenized by shaking at 1000 rpm, 1 minute. The concentrate was transferred to nuclease-free tubes and stored at 4°C and the RNA extraction was made on the same day.

### 2.3. RNA extraction

The RNA extraction was carried with Viral Nucleic Acid Extraction Kit II (GENEAID) with a modification: 200 µL of concentrated sample were homogenized and mixed with 20 µl of polyvinyl pyrrolidone 40 % (PVP) (Sigma) and 400 µl of lysis buffer, then homogenized 1 min and heated 10 min in dry block at 75 °C, followed by a 5 minutes centrifugation at 10,000 x g. The pellet was disposed and the supernatant was processed according to manufacturer’s instructions and RNA was eluted with 50 µl ultrapure water and stored at -20 °C until processing.

### 2.4. Viral detection and quantification

Viral RNA was detected by TaqMan 2019 nCOV Assay Kit v 1 (Applied Biosystems), in a Step One Plus (Applied Biosystems). TaqMan 2019nCoVControl Kit v1 (Applied Biosystems) was used as positive control.

The reaction mix (20 µl) consisted of: 6.25 µl TaqPathtm 1 step RTqPCR mastermix, GC; 1.25 µl of 2019 nCOV Assay (Gen ORF 1 ab) - FAM, 20X; 1.25 µl RNAse P-VIC, 20X; 11.25 µl of molecular biology quality water (Applied Biosystems).

The cycling conditions were as follows: UNG incubation: 25 °C, 2 minutes. Reverse transcription (RT): 50 °C, 15 minutes. Activation: 95 °C, 2 minutes. Cycling, 40 cycles including: Denaturation, 95 ° C, 3 seconds; Hybridization / Extension, 60 ° C, 30 seconds, collecting fluorescence data through FAM, VIC, HEX channels, using ROX as passive reference. The reaction volume was 25 µl. Each RNA obtained as well as the positive and negative controls were processed in duplicate.

### 2.5. Process control

As process control, Mengovirus (MGV) standard (CEERAMTOOLS®) described in ISO15216-1, 2017, was used [10]. Validation was carried out using 20 effluent samples collected over the course of two weeks under the same conditions as the samples to be analyzed. Once in the laboratory, 10 µl of MGV were inoculated to those samples, following the same concentration and extraction processes. The extracts obtained were also diluted 1/10.

The PCR and a MGV standard curve was performed according to the manufacturer’s instructions. The RT cycling conditions: 45 °C for 10 minutes, followed by a warm-up to95°C 10 minutes, and 45 amplification cycles of 15 seconds at 95 °C, followed by 45 seconds at 60 °C with acquisition of signal in this last stage. The FAM channel was used for fluorescence reading, using ROX as a passive reference.

The recovery rate of MGV was calculated following the guidelines established in the ISO 15216-1: 2017 [10].

## 3. Results and Discussion

The sampling extraction period was determined according to the highest measures of Escherichia coli concentrations. This range was defined between 9 am to 12 am. (Figure 2). PAC was chosen as an alternative option instead of AlCl3, widely indicated for concentration of enteric viruses in wastewater [11-12-13].

The RNA quantification was obtained by interpolating the quantification cycle (Ct) to a calibration curve from 5 duplicate serial dilutions of TaqMan 2019nCoVControl Kit v1 (10,000 copies/µl). The ORF 1ab calibration curve showed a linear range between 16 copies/µl to 10,000 copies/µl an amplification efficiency greater than 90% was taken as valid. A detection limit of 80 copies/µl (Ct value = 33, 91 ± 1.21).

The viral load was defined as the product between the concentration of copies/L of Orf 1ab gene, and the daily flow rate that entered to the plant on the day of the extraction. For this study, plants where associated with a served region, including medium to small sized plants with less than 420,000 equivalent inhabitants, and large-sized plants with 6.32 million equivalent inhabitants. Each treatment plant was considered separately, associating positive clinical cases[11][12] by served region.

For each plant, different linear regressions were performed, taking the log 10 of viral load as the independent variable, and the log 10 of positive cases as the dependent variable, considering the sum of positive cases for different time intervals.

Assuming the day of sample extraction as day zero, regressions were analyzed considering: Interval start from day -20 to day +18; Time intervals between 1 and 22 days of accumulated positive cases.

Significant correlations were observed by evaluating the accumulated positive cases across different periods of time respect a given sample date. For each WWTP different interval initial and central days were tested (Figure 3 to 10) and significant correlations were obtained by using 12 days intervals. The intervals of Table 2 which correspond to the correlation of Figure 11 were selected as representative.

**Figure 3.**
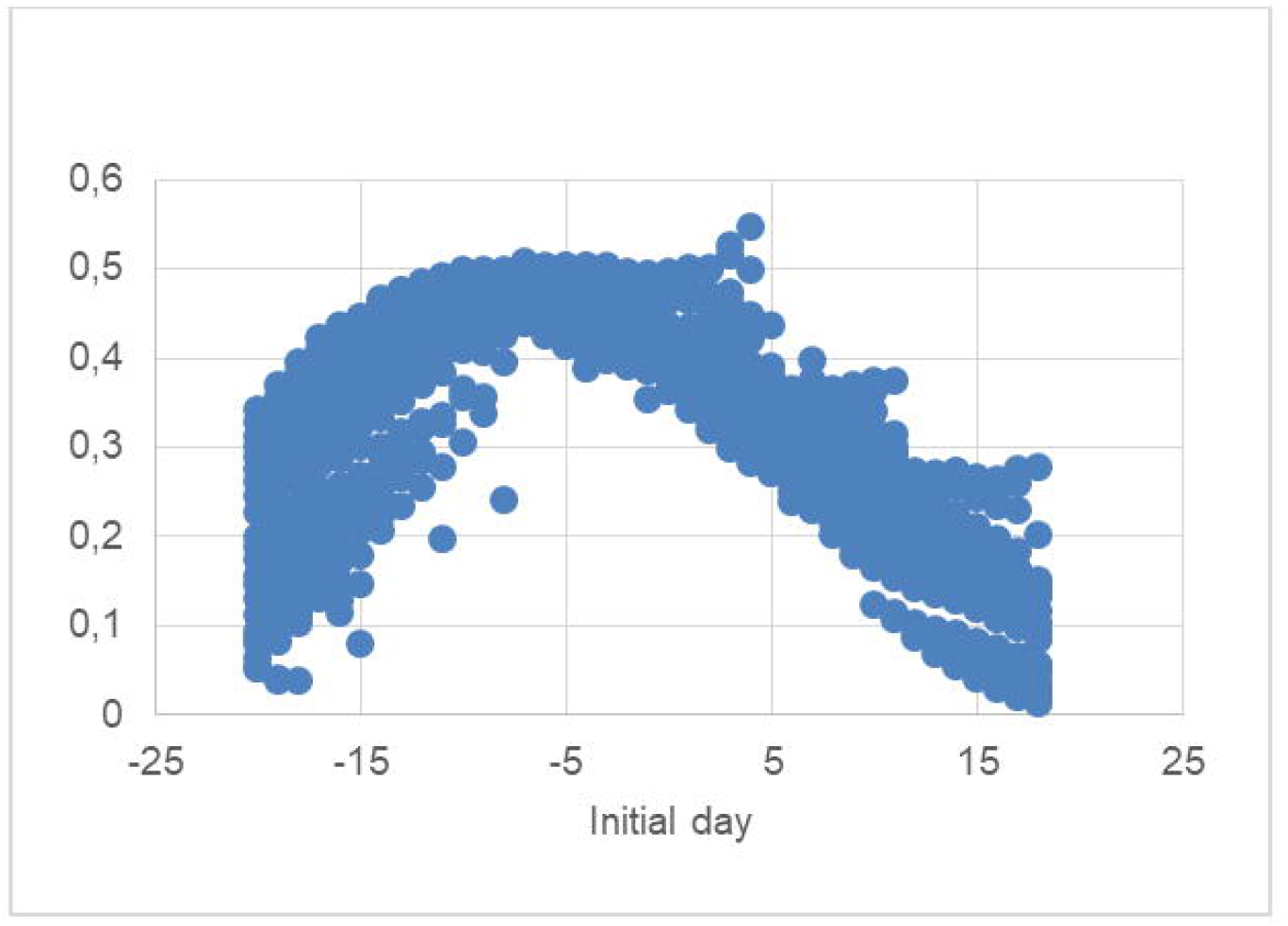
WWTP 1 - R^2^ vs Initial day.

**Figure 4.**
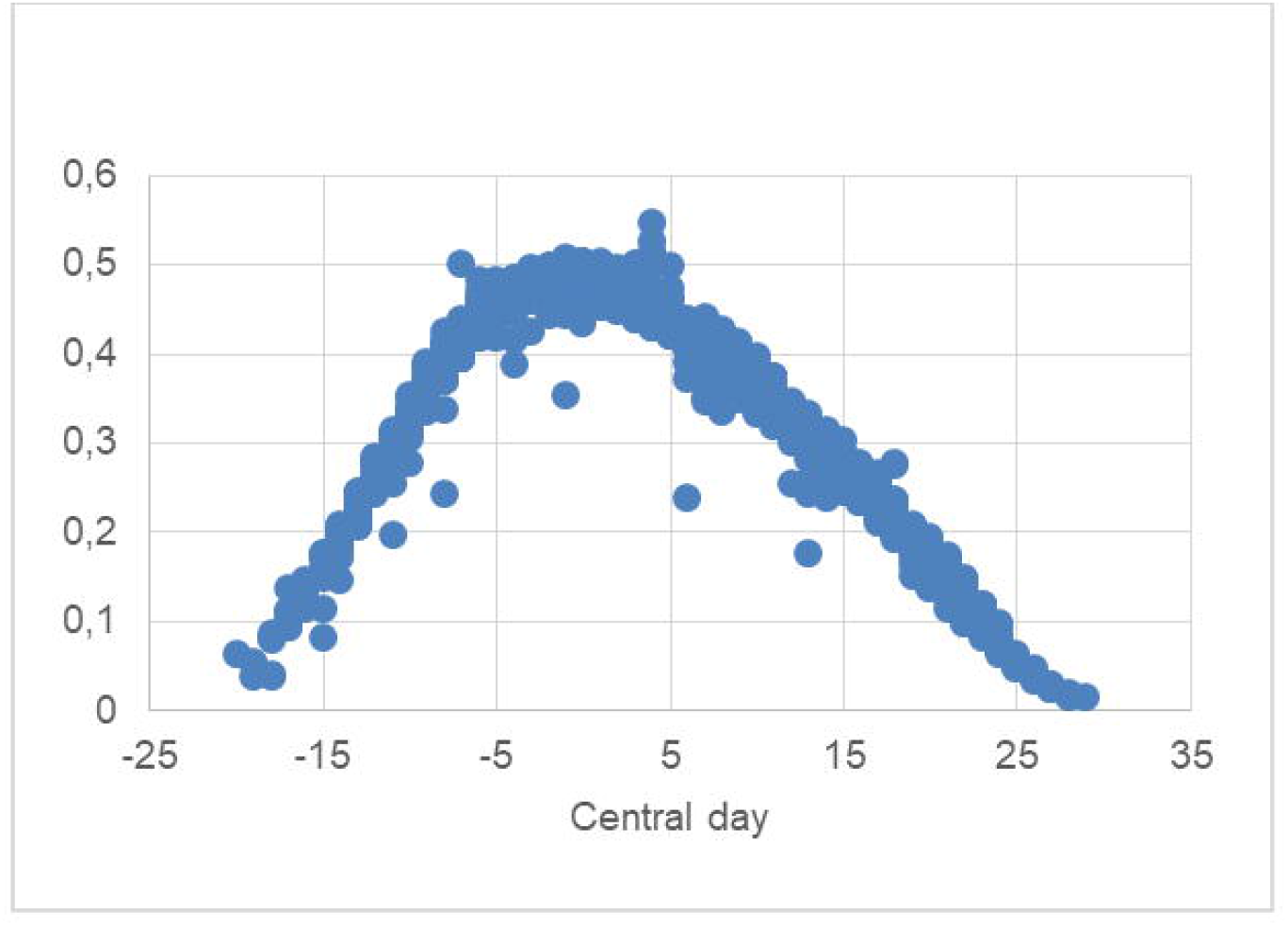
WWTP 1 - R^2^ vs Central day.

**Figure 5.**
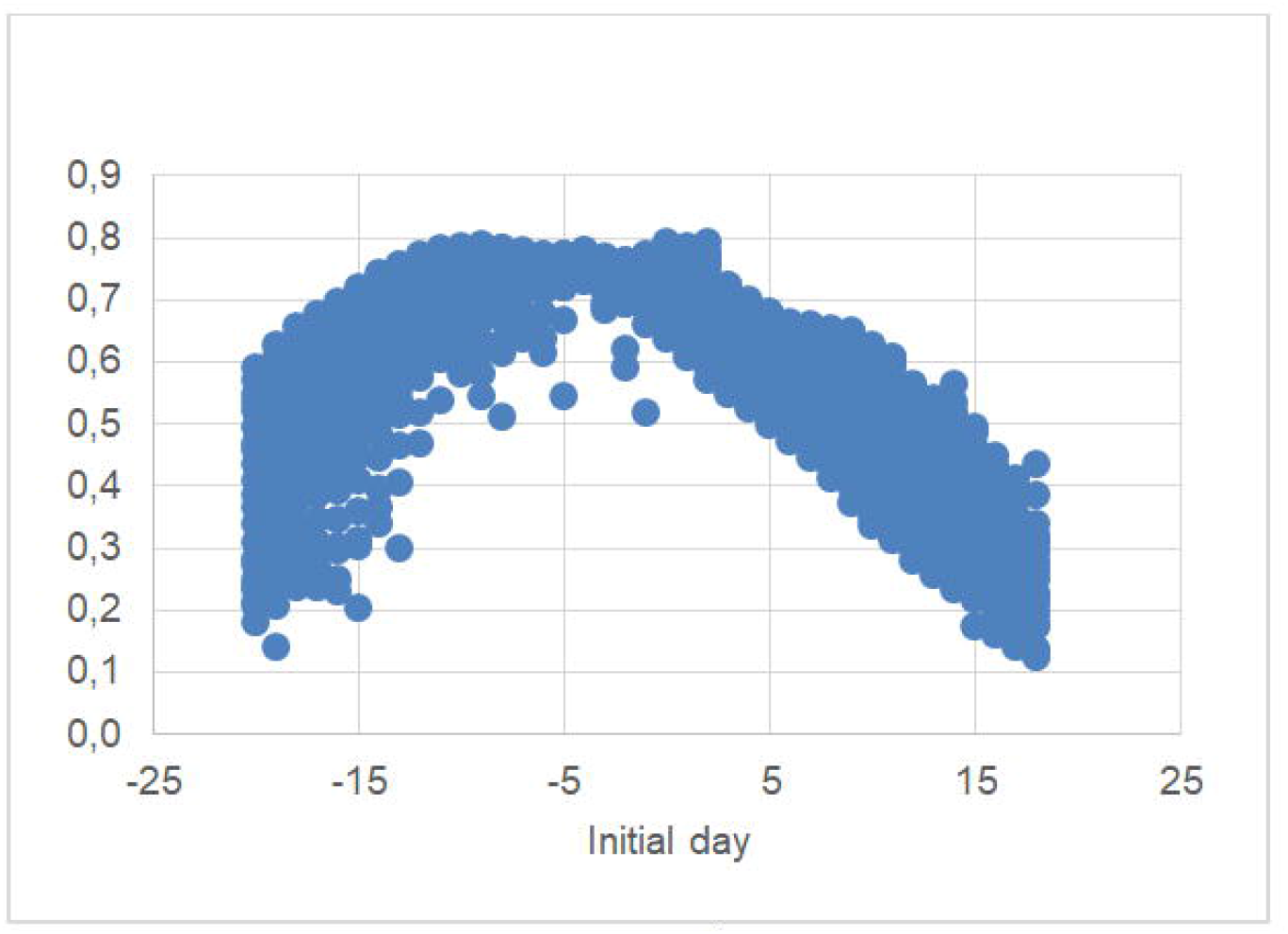
WWTP 2 - R^2^ vs Initial day.

**Figure 6.**
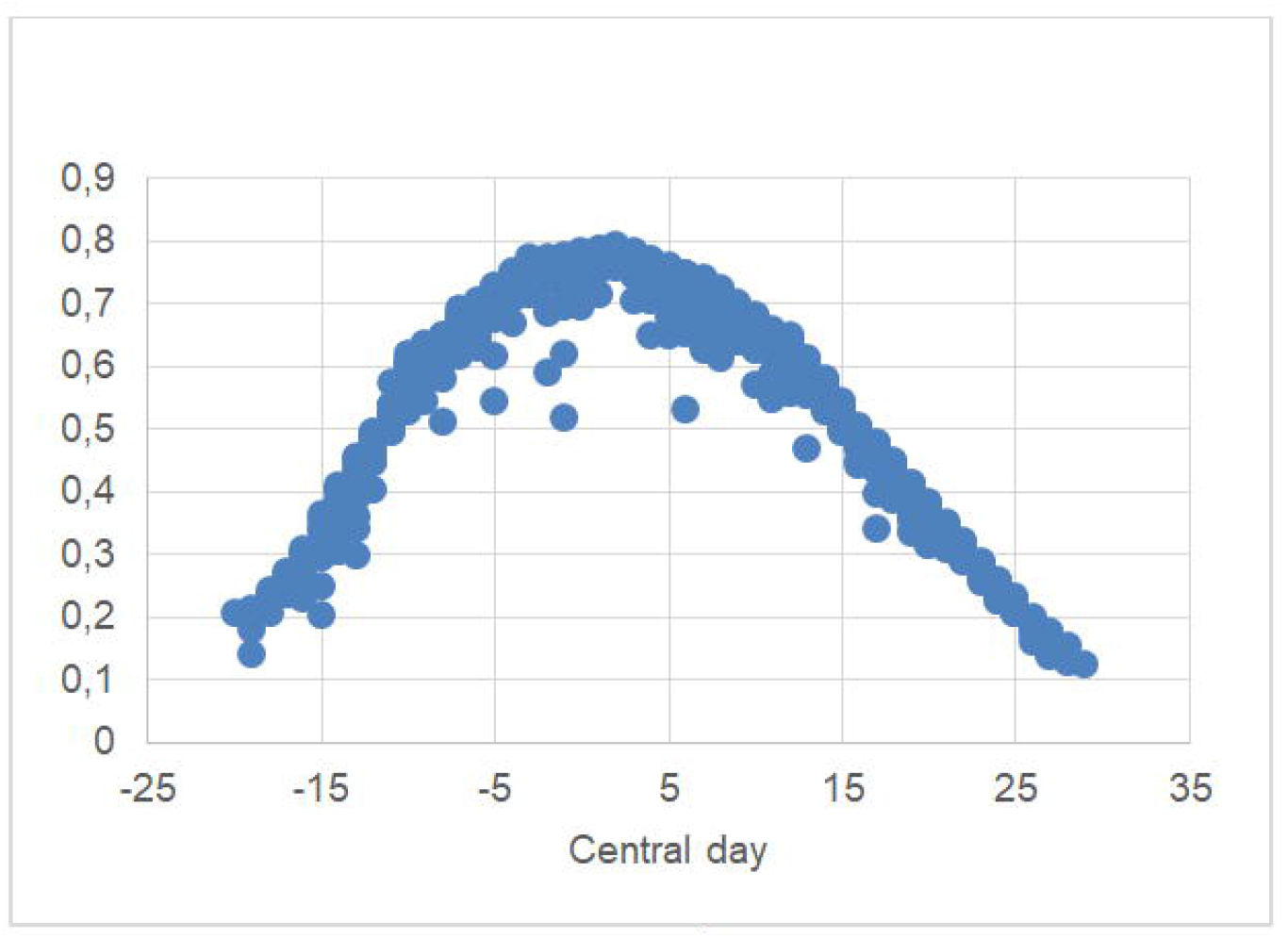
WWTP 2 - R^2^ vs Central day.

**Figure 7.**
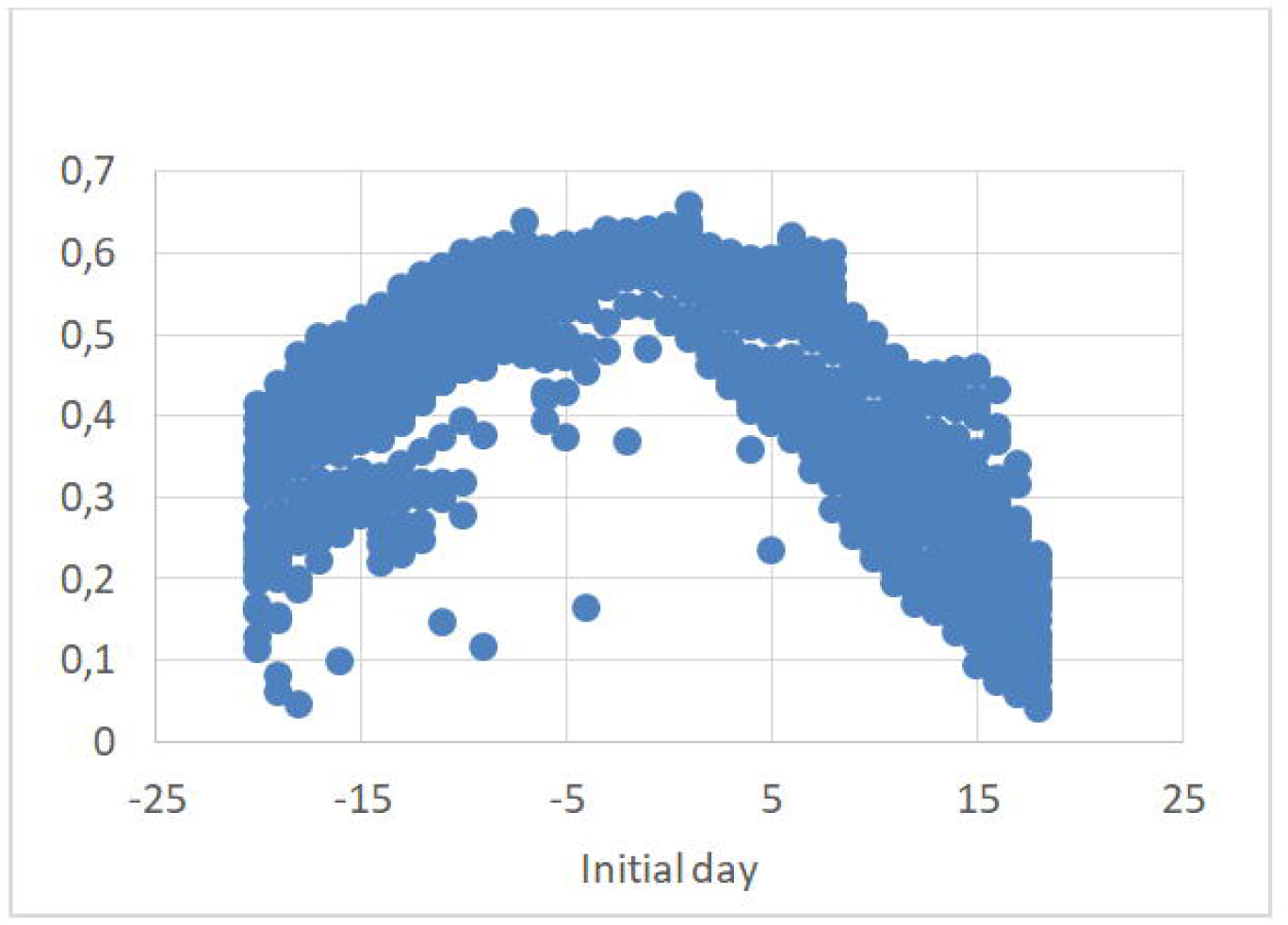
WWTP 3 - R^2^ vs Initial day.

**Figure 8.**
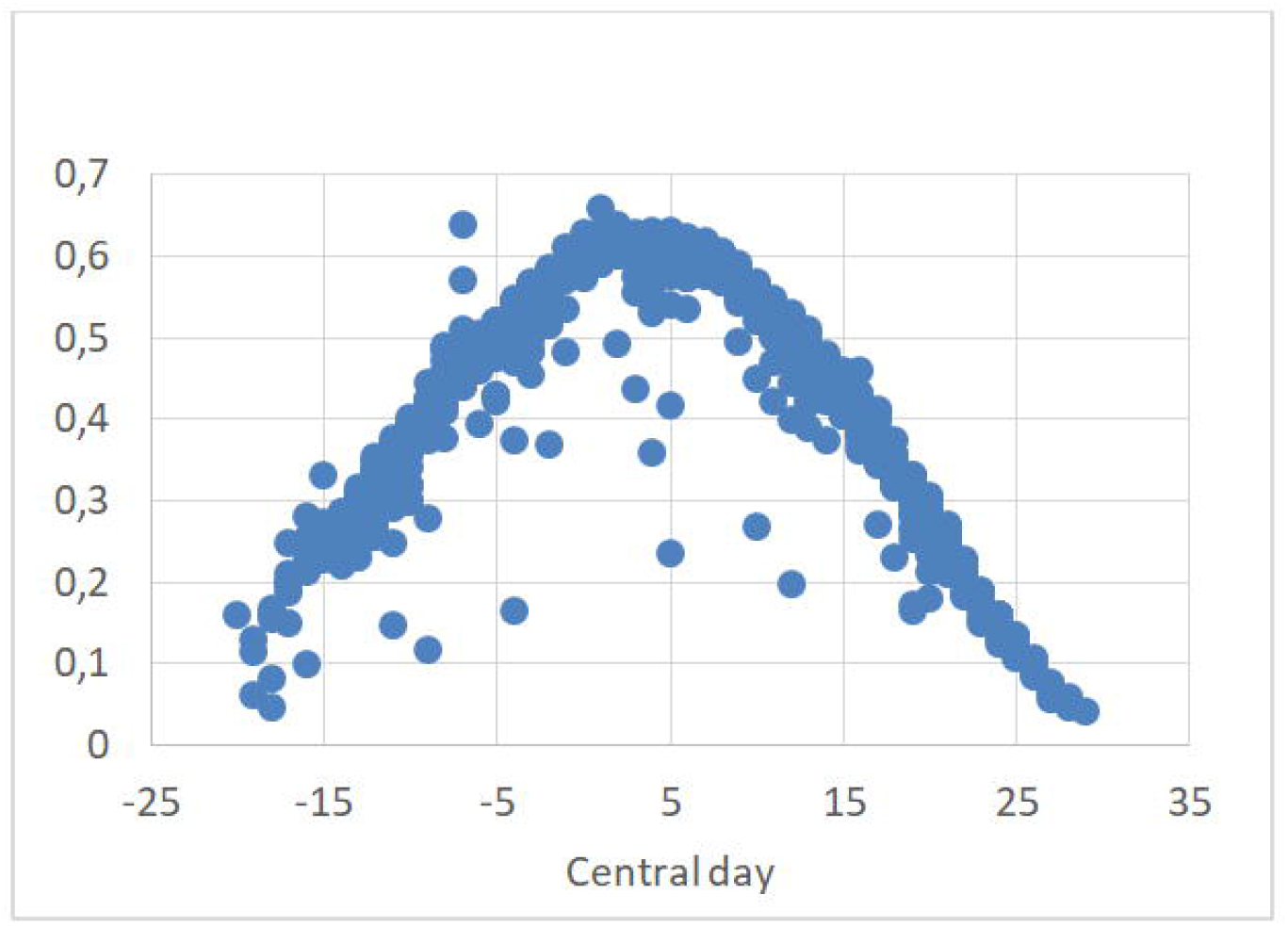
WWTP 3 - R^2^ vs Central day.

**Figure 9.**
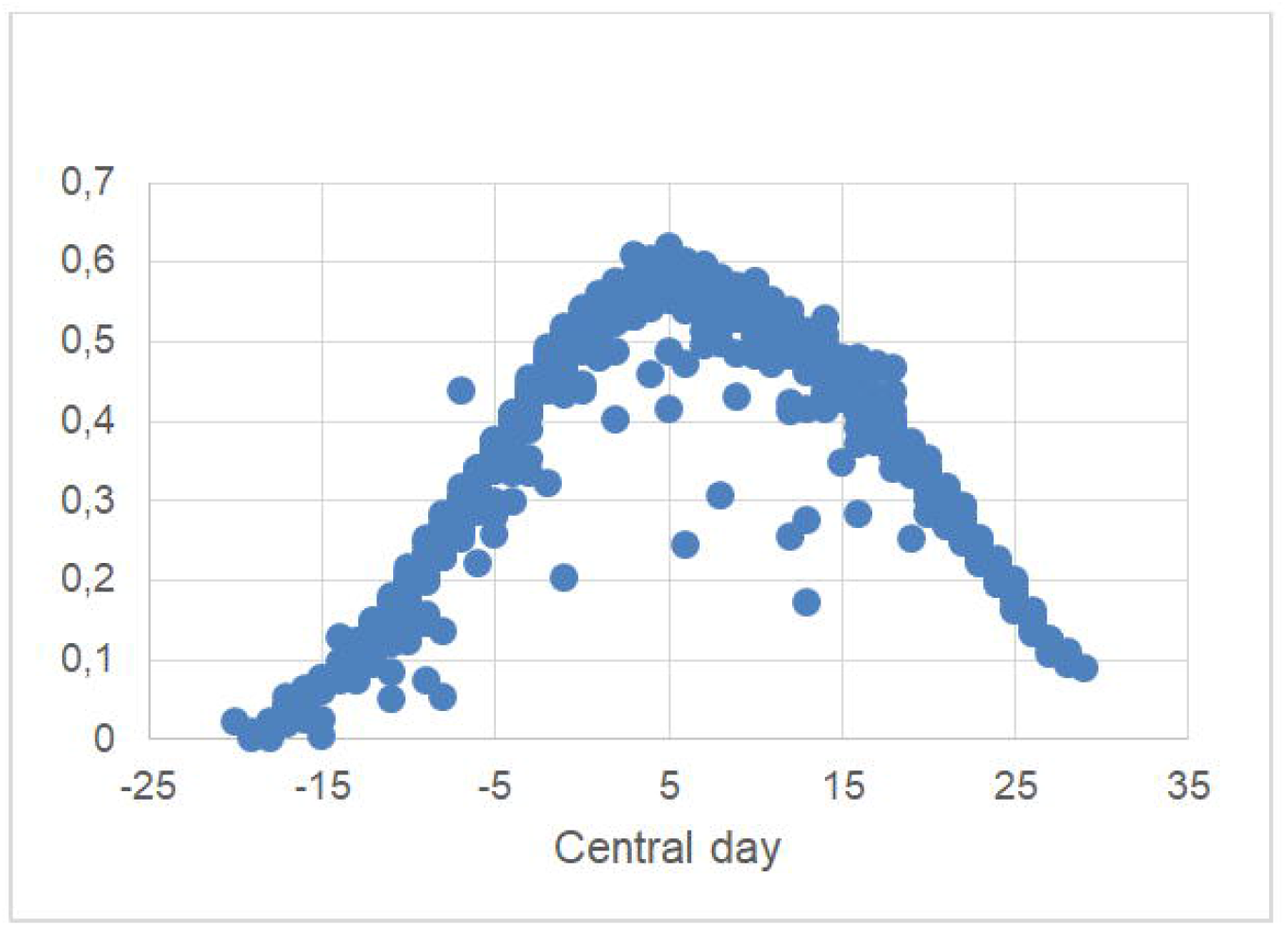
WWTP 4 - R^2^ vs Central day.

**Figure 10.**
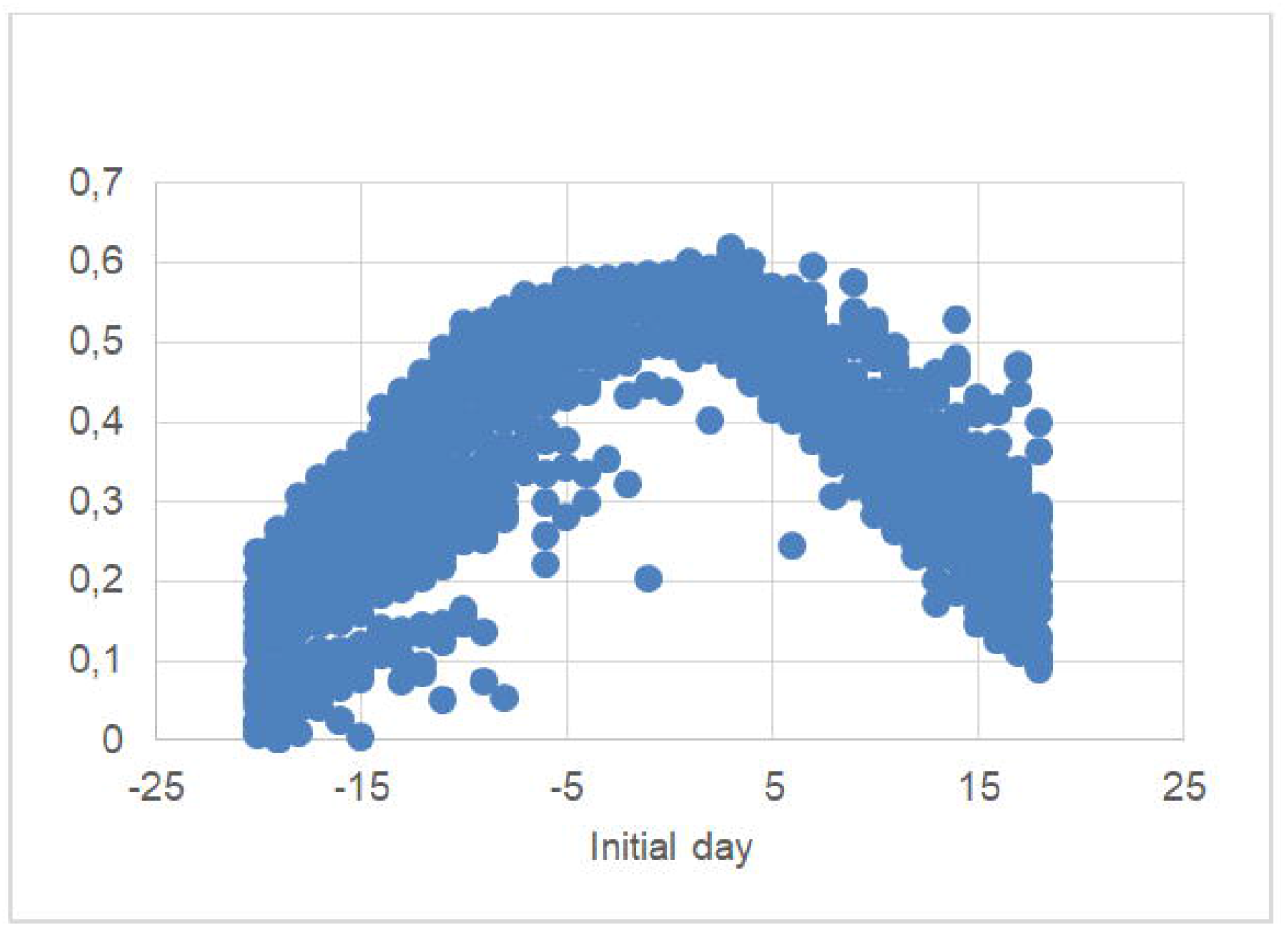
WWTP 4 - R2 VS Initial day.

**Table 2.** Representative intervals for each WWTP evaluated.

| | Initial day | Central day | $\Sigma$ days | $R^2$ | p-value |
| --- | --- | --- | --- | --- | --- |
| **WWTP 1** | -7 | -1 | 12 | 0,505 | 3,06E-04 |
| **WWTP 2** | -4 | 2 | 12 | 0,772 | 3,51E-07 |
| **WWTP 3** | -2 | 4 | 12 | 0,621 | 3,70E-05 |
| **WWTP 4** | -1 | 5 | 12 | 0,550 | 1,81E-04 |

**Figure 11.**
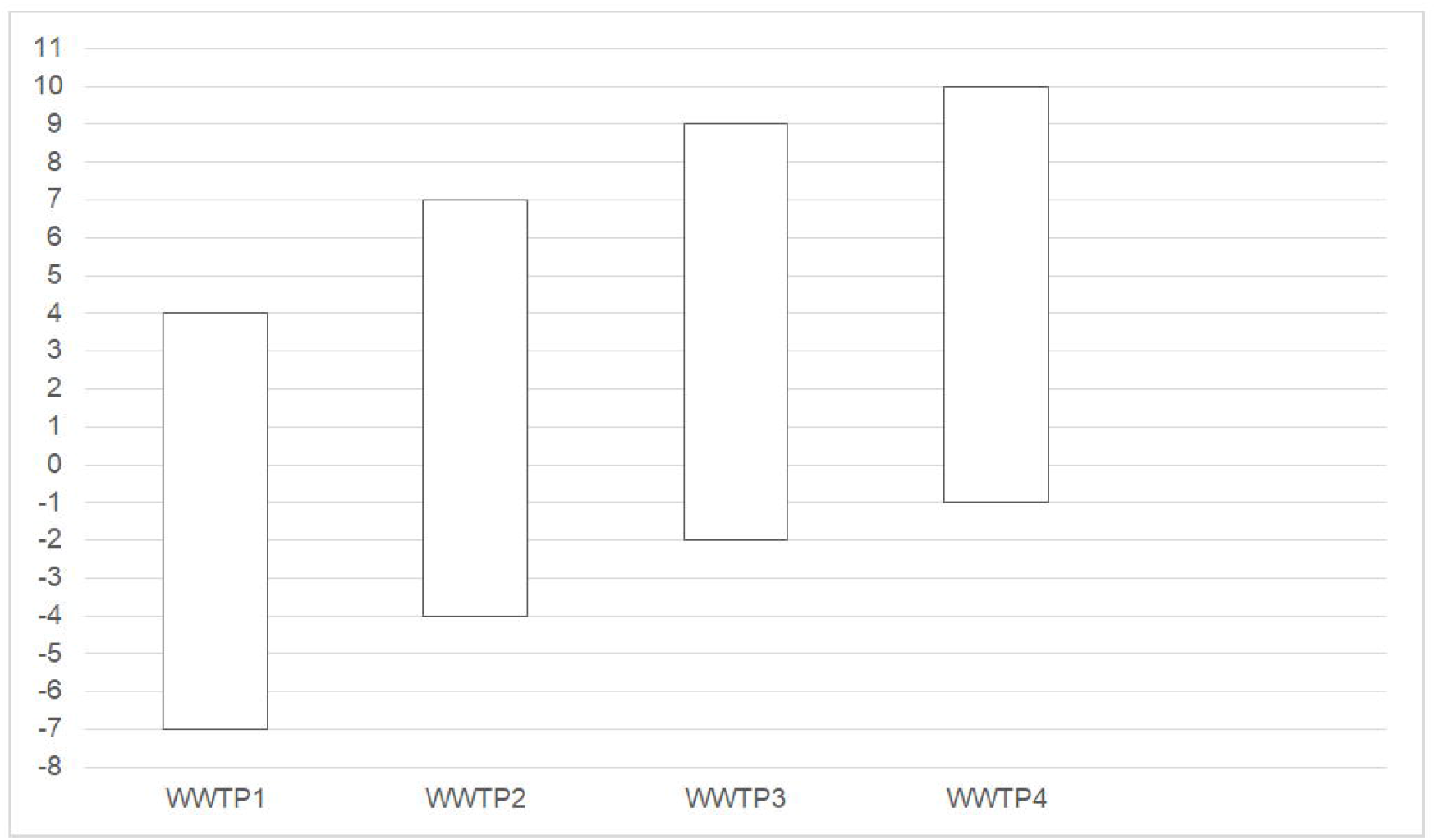
Descriptive scheme of representative intervals obtained on each WWTP.

In all cases, a similar behavior can be observed between the logarithms of the viral load and the positive cases reported (Figures 12,13,14,15).

**Figure 12.**
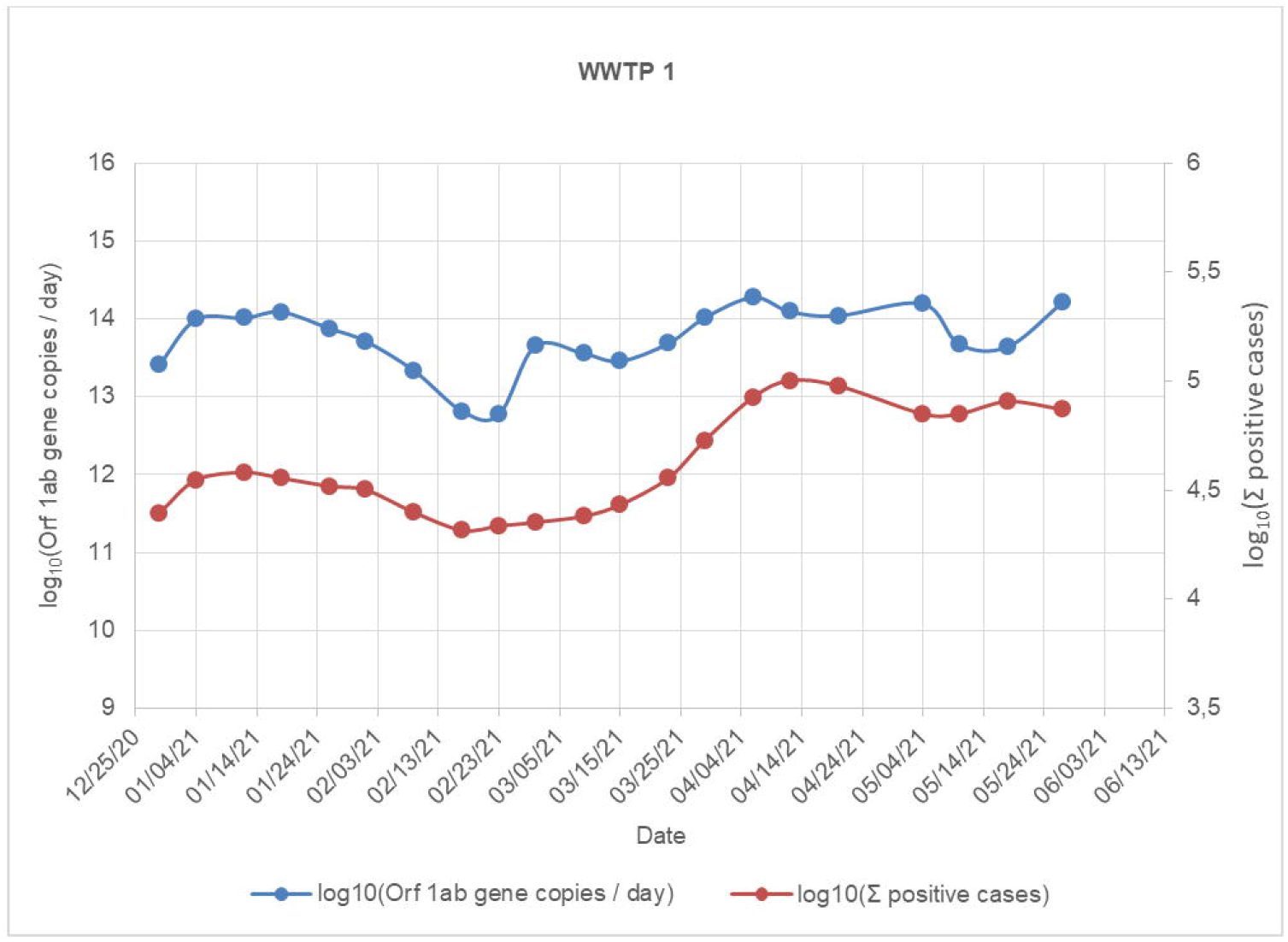
Evolution of Orf 1ab/day gene copies and accumulated positive cases (WWTP1).

**Figure 13.**
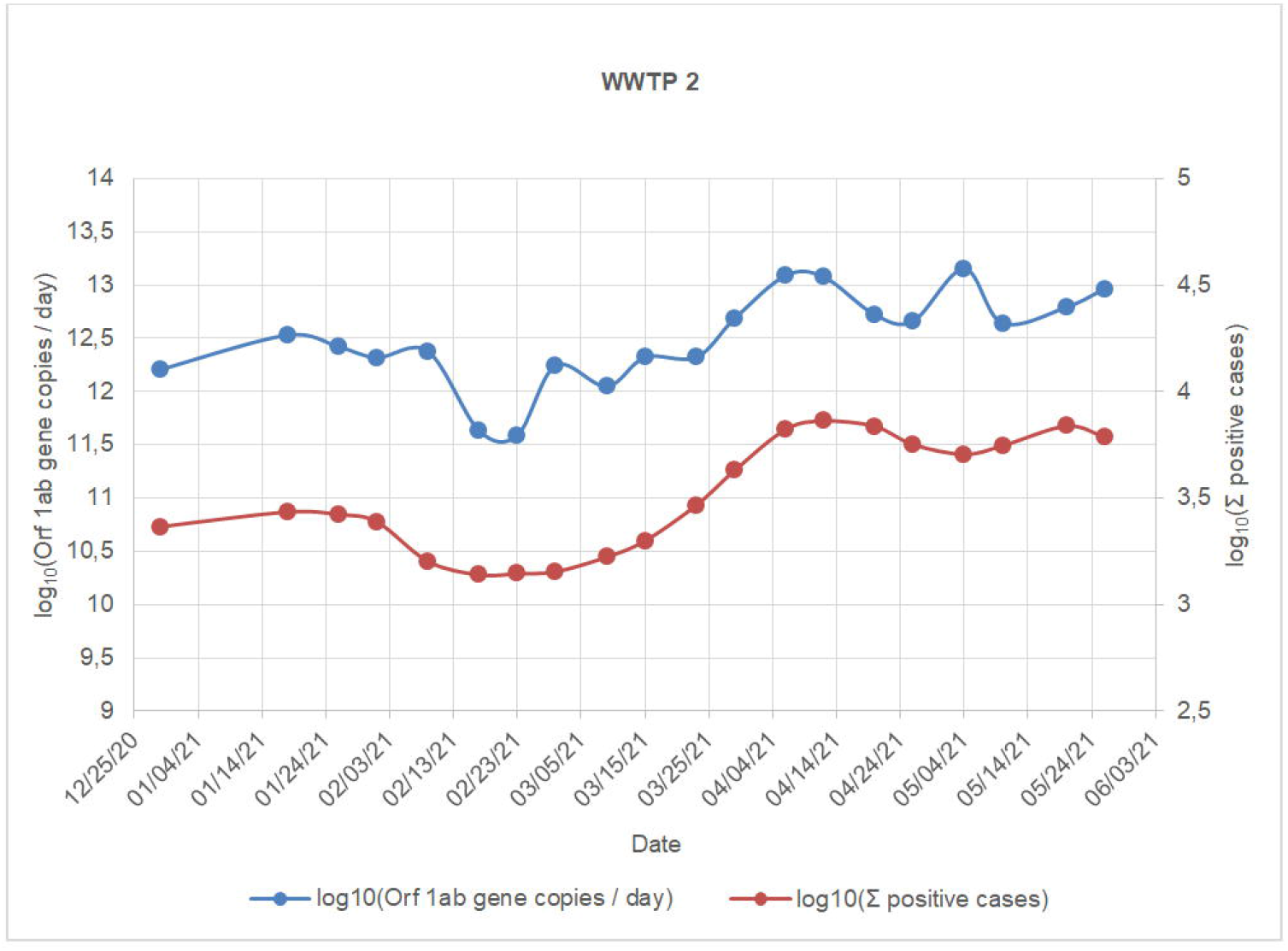
Evolution of Orf 1ab/day gene copies and accumulated positive cases (WWTP2).

**Figure 14.**
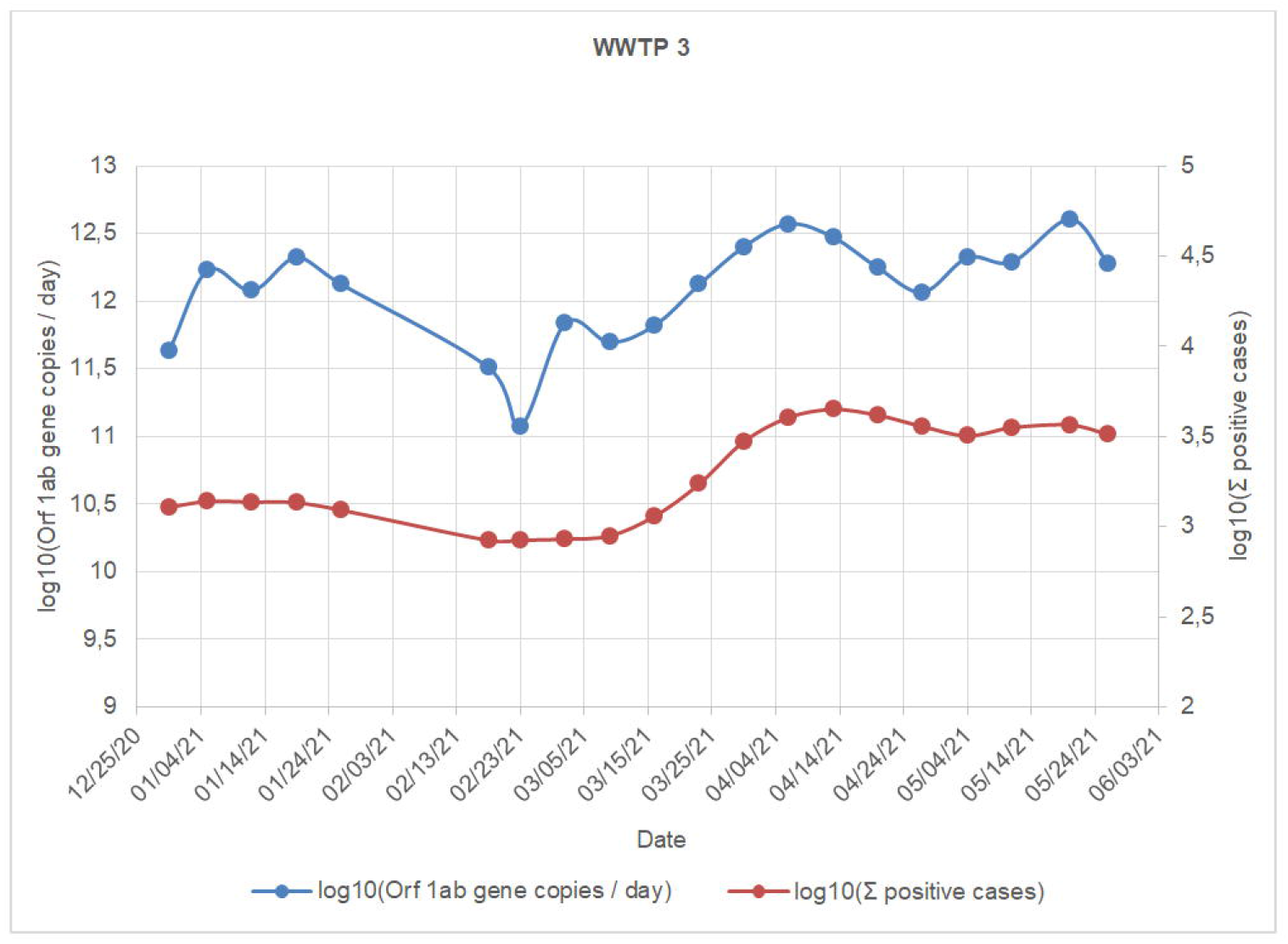
Evolution of Orf 1ab/day gene copies and accumulated positive cases (WWTP3).

**Figure 15.**
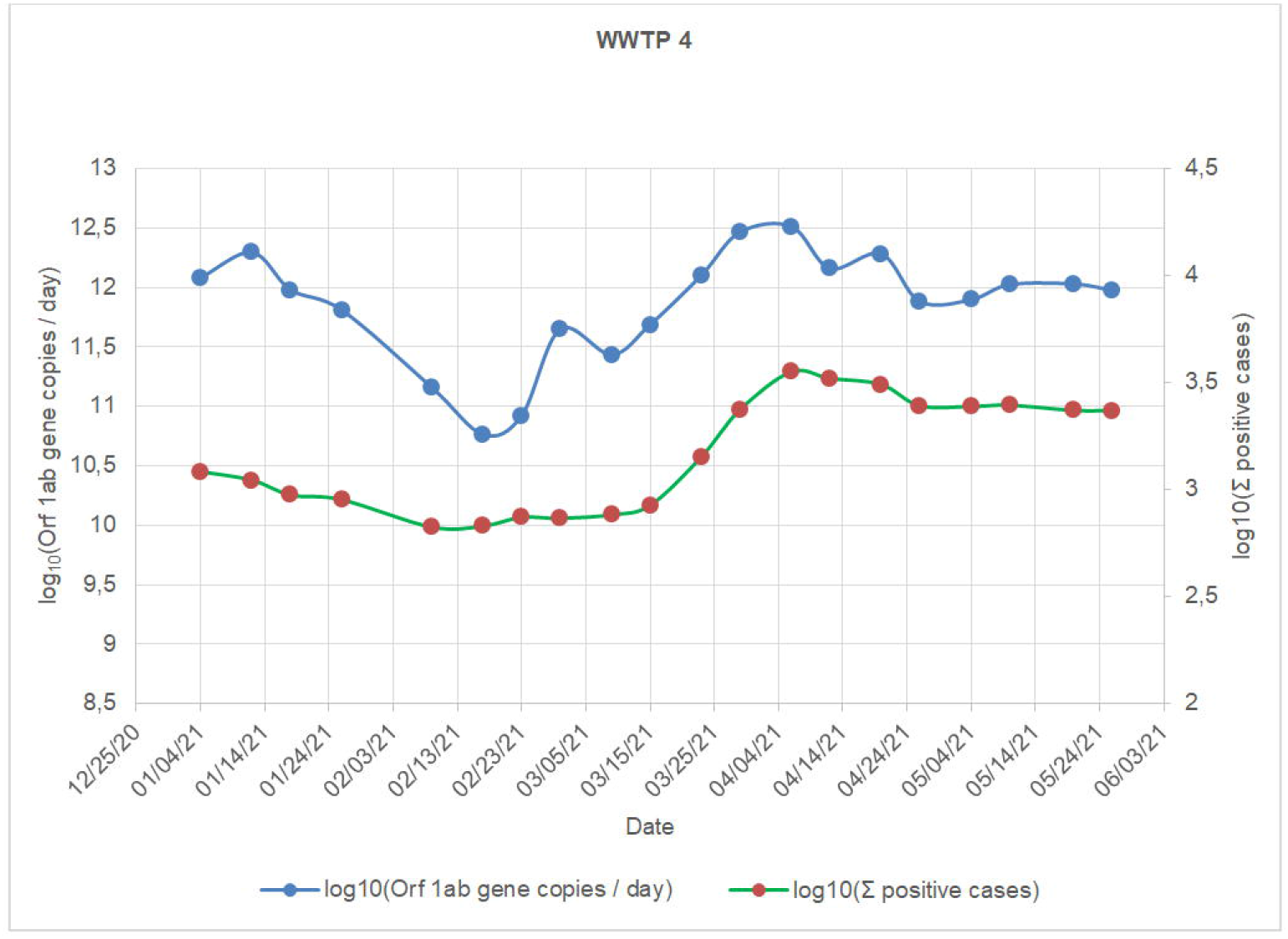
Evolution of Orf 1ab/day gene copies and accumulated positive cases (WWTP4).

An integral linear regression was tested for all plants, taking -4 as starting day and an interval of 12 days of accumulated positive cases for all plants. A correlation R2 = 0.8951, pvalue <2.2E-16, RSE = 0.2106 and adjusted R2 = 0.8937 (Figure 16), showed a satisfactory relationship which reinforces the idea of implementing the model in the region as a predictive tool.

**Figure 16.**
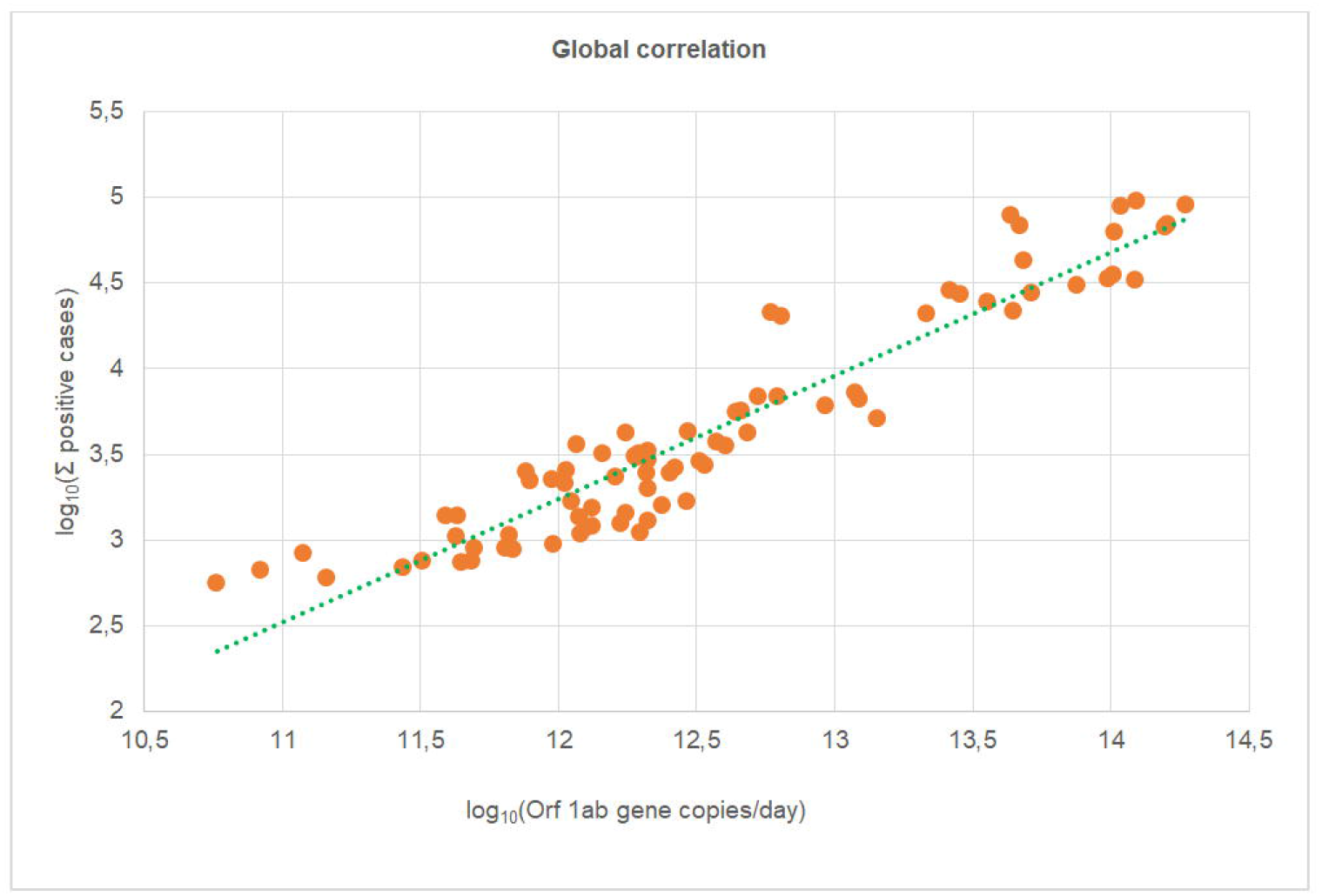
Global correlation (R2 = 0.8951, p-value < 2.2E-16).

The study covers the period of time between December 2020 to June 2021, when a peak of viral load was detected a week before the second wave of Covid-19. In this way, the method proved to be a useful and effective predictive tool.

With the aim to improve predictive aspects of this method, some variables such as temperature, hydraulic retention time, chemical composition of the matrix, flow rate variations, size and migration of the served population, clinical cases detection among others, should be considered in future studies.

## 4. Conclusions

The result of this initiative has been the early development of an integrated molecular sampling, analysis and detection system that can alert about the circulation of SARS-CoV-2 and offer valuable information on its prevalence in the population served to be used as a tool for epidemiological and environmental surveillance, as well as predict future waves of infection.

This development may be used for the analysis and control of other emerging pathogens that affect regions associated to a treatment plant.

## Supporting information

Correlations

## Data Availability

All data produced in the present study are available upon reasonable request to the authors

https://sisa.msal.gov.ar/datos/descargas/covid-19/files/Covid19Casos.zip

https://cdn.buenosaires.gob.ar/datosabiertos/datasets/salud/casos-covid-19/casos_covid19.csv

## Acknowledgements

We are grateful to Entidad Regional de Saneamiento y Depuración de Aguas Residuales (ESAMUR) for the technical assistance in the initial development of the molecular analysis methodology.

We are greatly thankful with Alejandro Branca BSc, for the invaluable support and contribution to this work.

